# Distinguishing Lewy Body Dementia from Alzheimer’s Disease using Machine Learning on Heterogeneous Data: A Feasibility Study

**DOI:** 10.1101/2022.04.10.22273665

**Authors:** Niamh McCombe, Alok Joshi, David P. Finn, Paula L. McClean, Gemma Roberts, John T. O’Brien, Alan J. Thomas, Joseph P.M. Kane, KongFatt Wong-Lin

## Abstract

Dementia with Lewy Bodies (DLB) is the second most common form of dementia, but diagnostic markers for DLB can be expensive and inaccessible, and many cases of DLB are undiagnosed. This work applies machine learning techniques to determine the feasibility of distinguishing DLB from Alzheimer’s Disease (AD) using heterogeneous data features. The Repeated Incremental Pruning to Produce Error Reduction (RIPPER) algorithm was first applied using a Leave-One-Out Cross-Validation protocol to a dataset comprising DLB and AD cases. Then, interpretable association rule-based diagnostic classifiers were obtained for distinguishing DLB from AD. The various diagnostic classifiers generated by this process had high accuracy over the whole dataset (mean accuracy of 94%). The mean accuracy in classifying their out-of-sample case was 80.5%. Every classifier generated consisted of very simple structure, each using 1-2 classification rules and 1-3 data features. As a group, the classifiers were heterogeneous and used several different data features. In particular, some of the classifiers used very simple and inexpensive diagnostic features, yet with high diagnostic accuracy. This work suggests that opportunities may exist for incorporating accessible diagnostic assessments while improving diagnostic rate for DLB.

**Clinical Relevance:** Simple and interpretable high-performing machine learning algorithms identified a variety of readily available clinical assessments for differential diagnosis of dementia, offering the opportunities to incorporate various simple and inexpensive screening tests for DLB and addressing the problem of DLB underdiagnosis.

## I. Introduction

Despite being the second most common form of neurodegenerative dementia, it is estimated that under half of dementia with Lewy bodies (DLB) cases are detected in routine clinical care [1, 2]. Accurate diagnosis, however, is critical in establishing accurate prognosis and comprehensive treatment plan [3].

Successive iterations of clinical DLB criteria have become more sensitive and less specific over time, particularly as biomarkers such as ^123^I-2β-carbomethoxy-3β-(4-iodophenyl)-N-(3-fluoropropyl) nortropane SPECT (FP-CIT), and ^123^I-metaiodobenzylguanidine (MIBG) cardiac scintigraphy have emerged [4]. However, these investigations are not available to all clinical services, and may be infeasible in some patients. It is therefore important that opportunities to characterise the clinical features (i.e., the signs and symptoms identified by the healthcare practitioner) are pursued.

Machine learning (ML) methods [5] may offer one such approach to clinically characterizing DLB. Identification of subphenotypes using ML have assisted clinical decision-making in Alzheimer’s disease (most common form of dementia) [6], and disorders such as delirium [7] and sporadic Creutzfeldt-Jakob disease [8]. In contrast, most DLB studies to date have adopted traditional statistical methods to provide group-level insights that may not apply to all subgroups within DLB. Recent studies applying ML, particularly, deep learning on structural or functional neuroimaging data could detect DLB from AD with accuracy in the range ∼71 – 89% [9-11]. However, these ML algorithms were complex and not readily interpretable and were not applied to heterogeneous data, including readily accessible assessments.

This work aims to determine the feasibility of using simple and interpretable machine learning on heterogeneous data, including accessible diagnostic assessments, of well characterised DLB and AD research cohorts to identify novel subphenotypes based on clinical observations alone.

## II. methods

### A. Data Description

Data was collected as part of a study investigating the diagnostic utility of MIBG and FP-CIT in a clinically representative UK cohort [12]. Participants met research criteria for either probable DLB [13] or probable AD [14]. Subjects were all over 60 years old at recruitment, and recruited through psychiatry of Old Age, geriatric medicine and neurology clinics in North-East England between September 2015 and June 2017. Written informed consent was provided by patients, or by a nominated consultee when participants did not demonstrate capacity to consent for themselves. Each subject underwent detailed clinical examination that included multiple rating scales and neuropsychological testing, data from which were used for this study.

### B. Data Pre-processing

The resultant dataset had 233 data feature columns including the Diagnosis column, with 33 rows representing cases. The Diagnosis column had 17 cases diagnosed with DLB and 16 cases diagnosed with AD and was used as the outcome variable. This dataset had 7% missing values, which were imputed, separately from the Diagnosis column to avoid “double-dipping” [15], using the missForest algorithm [16] and R package [17]. The missForest algorithm was used as previous work by the authors found it to be an accurate and flexible imputation algorithm [18].

### C. Random Forest Feature Importance

To obtain an overview of the most relevant variables, a Random Forest (RF) model [19] was built using the whole imputed dataset, and the randomForest package in R [20]. The variable importance was extracted from the model and plotted along a single-dimensional plot using the lattice package [21]. FP-CIT and MIBG related features were highlighted separately in the plot, as they are excluded from some of the analysis.

### D. Association Rule-Based Classifiers

A Leave-One-Out-Cross-Validation (LOOCV) technique, in which each row of the dataset was omitted in turn, was used to extract 33 association rule-based classifiers from the data using the Repeated Incremental Pruning To Reduce Error Production (RIPPER) algorithm [22] as implemented in the RWeka package [23]. This simple classifier was selected for two reasons: (i) to avoid overfitting given that the data is high-dimensional with a small sample size; and (ii) to maximize the transparency of interpretation of the algorithm to encourage clinical adoption.

The dataset was then pruned to remove data related to FP-CIT and MIBG, as the goal is to identify novel clinical diagnostic DLB subphenotypes. The LOOCV process was run again on this reduced dataset and 33 new rule-based classifiers were extracted by RIPPER.

In the case where any of the rule-based classifiers extracted on different iterations of LOOCV were identical to each other, the total number of instances of each rule-based classifier was recorded. Each classifier was then tested on the entire dataset, and class-balanced accuracy, sensitivity and specificity (with DLB as the positive class) was recorded.

Each classifier was separately used to classify the omitted individual case (the latter not being used in the training dataset when the classifier was built), and the total number of correct classifications of omitted subjects was recorded.

### E. Testing statistical validity of extracted rules

Because this dataset is high-dimensional, with far more features than cases, it is important to guard against the risk of overfitting (when a model does not generalize beyond the training data). It could be speculated that with 232 features to choose from, the RIPPER algorithm will always find some diagnostic rules that can accurately classify the 33 cases. As a precaution against this, the methodology was tested against noise data with the same dimensionality and variable distributions but randomly scrambled from the original dataset. Specifically, the values of the dataset features were permuted (i.e. the columns of the dataset were shuffled) using the modelr package [24] in R. 100 permuted datasets were generated and 100 rules-based classifiers were extracted from these datasets using RIPPER. A simple t-test was used to determine whether the overall accuracy of classifiers built on permuted datasets in classifying their (permuted) training data was significantly different from the overall accuracy of the classifiers built on the original dataset, in classifying their training data. A significant difference here would suggest that the algorithm is detecting valid rather than spurious patterns in the original data.

### F. Software and Hardware for Analysis

The above analyses and algorithms were run within R Studio version 1.146 on a Windows machine with eight memory cores, Intel i7 processor, 16GB RAM and R version 3.5.2 installed. Source codes for the computations are available at https://github.com/mac-n/DLB. The dataset used during the study is available from the corresponding author (JPMK) on reasonable request.

## III. results

### A. Ranking Feature Importance Identified Heterogeneous Set of Assessments

A strip plot of the Random Forest Importance (RFI), as determined by decrease in Gini index [25], of all the data features is shown in Fig. 1, and the names of the top-ranked features in Table I (described in Appendix Table I). Data features deriving from FP-CIT and MIBG are coloured in red in Fig. 1 and are in bold text in Table I. It can be seen in Fig. 1 that the vast majority of the 232 features sit at the left of the plot, meaning very few features had a strong impact on the classifier accuracy (for DLB-vs-AD diagnosis). Other than the Trail Making Test – Part A [26] (trailsA), the other top-ranked 14 features in Table I were expected as they are currently directly or indirectly contributing to the diagnosis of DLB. Hence, it was interesting to see the algorithm identifying a new feature for potential assessment.

**Figure 1.**
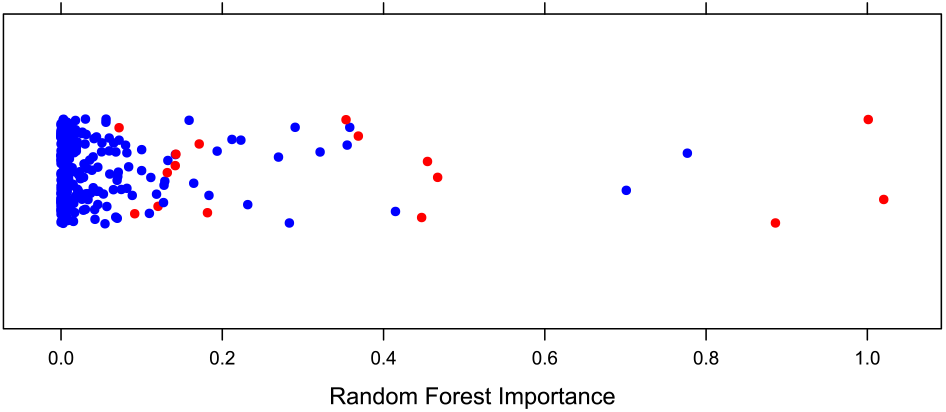
Strip plot of Random Forest Importance (RFI) of all features in the dataset. See Table I for these specific features. Red dots: Features deriving from FP-CIT or MIBG.

**TABLE 1.**
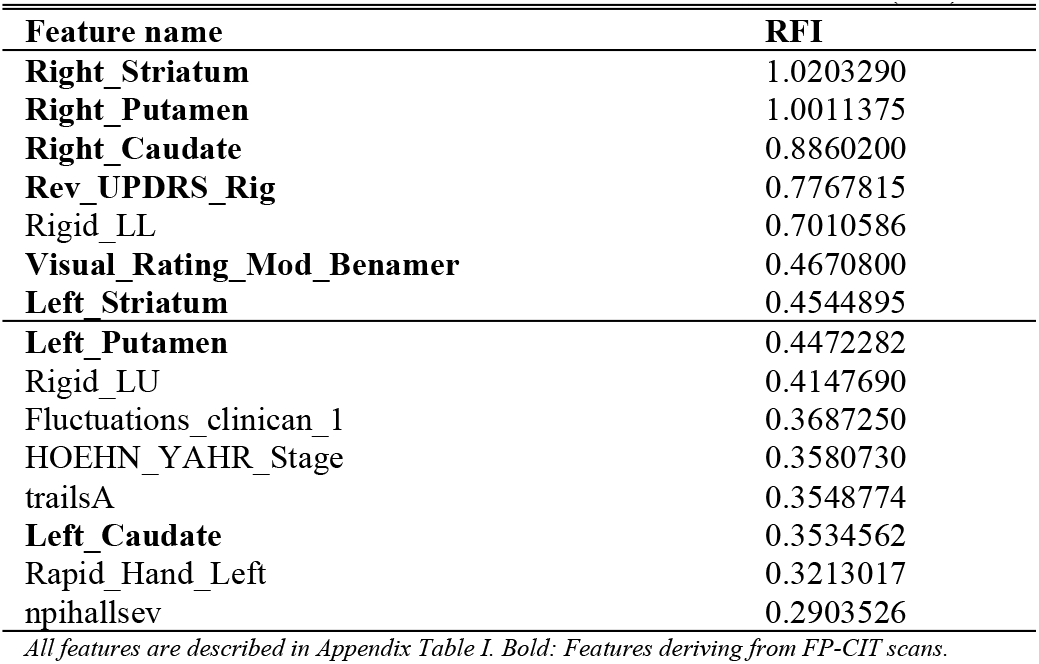
Top Features Ranked By Random Forest Importance (RFI)

### B. Simple, Readily Available Assessments with Classification Accuracy

Table II shows all the association rule-based classifiers selected with this dataset using the RIPPER algorithm and LOOCV, and their classification accuracy over the whole dataset. The classifier rules shown in the table are simple to interpret. Specifically, the rules can be understood as stating that if the condition on the left of ⇒is true, the outcome value on the right should be predicted. The classifier follows each rule in the order listed. If none of the rules apply, the classifier chooses a default value.

It can be seen that the Unified Parkinson’s Disease Rating Scale (UPDRS) [27] - Rigidity subscale (Rev_UPDRS_Rig) was most frequently selected as the basis of classification. This relates to the impairment of movement often associated with DLB [28]. FP-CIT brain imaging variables were also frequently selected for the classifier. This was expected, given the important role of FP-CIT in influencing the clinical diagnosis of DLB [4]. Thus, this validated the use of the association rule-based classifier.

**TABLE II.** Association Rule-Based Classifiers Selected By RIPPER with LOOCV

| Rules | # | Acc | Sens | Spec |
| --- | --- | --- | --- | --- |
| (Rev_UPDRS_Rig $\leq 0$) ⇒<br>Final_diag=AD | | | | |
| otherwise Final_diag=DLB | 22 | 0.94 | 0.88 | 1 |
| (Rigid_LL $\leq 0$) ⇒ Final_diag=AD | | | | |
| otherwise Final_diag=DLB | 3 | 0.9 | 0.8 | 1 |
| (Left_Striatum $\geq 1.4415$) and (Rigid_LL $\leq 0$) ⇒ Final_diag=AD | | | | |
| otherwise Final_diag=DLB | 2 | 1 | 1 | 1 |
| ((Right_Striatum $\geq 1.55$) ⇒<br>Final_diag=AD | | | | |
| otherwise Final_diag=DLB | 2 | 0.94 | 0.94 | 0.94 |
| (Rigid_LU $\leq 0.07$) ⇒ Final_diag=AD | | | | |
| otherwise Final_diag=DLB | 2 | 0.87 | 0.76 | 1 |
| (DCFS_drowsy $\leq 2$) and (Rigid_RU $\leq 0$) ⇒ Final_diag=AD | | | | |
| otherwise Final_diag=DLB | 1 | 0.94 | 0.94 | 0.94 |
| (Right_Caudate $\geq 1.491$) ⇒<br>Final_diag=AD | | | | |
| otherwise Final_diag=DLB | 1 | 0.9 | 0.8 | 1 |
| (Left_Caudate $\geq 1.62$) ⇒<br>Final_diag=AD | | | | |
| otherwise Final_diag=DLB | 1 | 0.9 | 0.88 | 0.93 |

**Rules:** Ruleset describing the classifier
**#:** Number of times classifier was mined with RIPPER using LOOCV
**Acc:** Whole-sample accuracy of classifier
**Sens:** Whole-sample sensitivity of classifier (positive class: DLB)
**Spec:** Whole-sample specificity of classifier

Each classifier was then tested on the out-of-sample subject which was not used in this training data. The mean accuracy was 76% i.e., 25 of the 32 classifiers predicted the correct diagnosis for their out-of-sample patient. Mean sensitivity was 0.9 and mean specificity was 0.68, with DLB as the positive class.

Next, we aim to identify more readily accessible assessments. Table III shows the association rule-based classifiers selected when FP-CIT and MIBG features were removed. Even with this, the classification accuracy remained very high, as discussed below. We can also observe in Table III that the UPDRS Rigidity subscale is still frequently selected as the sole basis for classification, and in one instance, combined with the Neuropsychiatric Inventory (NPI) [29] – Hallucinations subscale. Interestingly, specific rigidity scores within this UPDRS Rigidity subscale such as Rigid_LL (left leg rigidity) constituted accurate diagnostic classifiers on their own. Moreover, in both Tables II and III, the selected rule-based classifiers were all very simple, consisting of only one or two rules for diagnosis. Note that since these classifiers produced highly accurate performance on the training data, the RIPPER algorithm did not seek out additional rules.

**TABLE III.** Association Rule-Based Classifiers Selected By RIPPER with LOOCV without FP-CIT and MBIG

| Rules | # | Acc | Sens | Spec |
| --- | --- | --- | --- | --- |
| Rigid_LL $\geq 1$) ⇒ Final_diag=DLB | 10 | 0.97 | 0.94 | 1 |
| (Rey_ListB $\leq 1$) ⇒ Final_diag=DLB | | | | |
| otherwise Final_diag=DLB |  |  |  |  |
| (Rev_UPDRS_Rig $\leq 0$) ⇒ Final_diag=AD | 9 | 0.94 | 0.88 | 1 |
| otherwise Final_diag=DLB |  |  |  |  |
| DCFS_Sleep $\leq 3$) and (Rigid_LU $\leq 0$) ⇒<br>Final_diag=AD | 5 | 1 | 1 | 1 |
| otherwise Final_diag=DLB |  |  |  |  |
| (Rev_UPDRS_Rig $\geq 1$) ⇒<br>Final_diag=DLB | 4 | 0.94 | 1 | 0.88 |
| otherwise Final_diag=AD |  |  |  |  |
| ((npihall $\geq 0.8$) ⇒ Final_diag=DLB | 2 | 1 | 1 | 1 |
| (Rev_UPDRS_Rig $\geq 2$) ⇒<br>Final_diag=DLB | | | | |
| otherwise Final_diag=AD |  |  |  |  |
| (Rigid_LL $\leq 0$) ⇒ Final_diag=AD | 1 | 0.97 | 0.94 | 1 |
| otherwise Final_diag=DLB |  |  |  |  |
| ((UPDRS_Total $\leq 7$) ⇒ Final_diag=AD | 1 | 0.94 | 0.94 | 0.94 |
| (ACE_recog $\leq 1$) ⇒ Final_diag=AD | | | | |
| otherwise Final_diag=DLB |  |  |  |  |

**Rules:** Ruleset describing the classifier
**#:** Number of times classifier was mined with RIPPER using LOOCV
**Acc:** Whole-sample accuracy of classifier
**Sens:** Whole-sample sensitivity of classifier (positive class: DLB)
**Spec:** Whole-sample specificity of classifier

When these association rule-based classifiers were each tested in turn on the out-of-sample subject which was not used in their training data, mean accuracy was 85% (95% CI 0.681, 0.9489) i.e., 28 of the 33 classifiers predicted the correct diagnosis for their out-of-sample patient. Mean sensitivity was 0.88 and mean specificity was 0.82, with DLB as the positive class. Given the very simple nature of the classifiers, this is a strong performance on the out-of-sample test data.

Results in both Table II and Table III suggest possible diagnostic screening routes that do not involve FP-CIT or MIBG but include some very simple and readily available assessments. The identified features other than FP-CIT features were Rev_UPDRS_Rig, Rigid_LL, npihall, DCFS_Sleep, DCFS_Drowsy, Rigid_LU, UPDRS_Total, ACE_recog, and Rey_ListB. These features are described in Appendix Table 1. It should be noted that these data features, even when used individually or in pairs for diagnosis as is the case here, can produce highly accurate diagnostic classifiers (≥ 94%). It is also interesting to note that a wide variety of data features can be used for these accurate diagnostic predictions; this suggests that even if some of these features are later eliminated as potential candidates for use in DLB diagnosis after more extensive testing, or unavailable in certain clinics, many other promising avenues of assessments remain.

### C. Validation of Results

To verify that the classifiers selected by RIPPER were not the result of overfitting on this low-N high-dimensional dataset, the methodology was tested on 100 noise-generated datasets with the same dimensionality (Section II.E). The accuracy of each classifier in classifying its noise-generated training dataset was compared with the accuracy of classifiers shown in Tables II and III. The mean accuracy of the 100 classifiers trained on noise-generated data when classifying their training data was 78%, with varying performance ranging from 51% to 100%. By contrast, the mean accuracy of the classifiers described in Table II and Table III was 94% when classifying their training dataset, with performance ranging from 90% to 100%. A simple t-test determined the classifiers trained on noise-generated data to perform significantly different from the classifiers in Tables II and III, (p<2 x10^−16^). Thus, this demonstrated that our results (Tables II and III) were not the product of overfitting.

## IV. discussion

Our previous work [12] had used receiver operating characteristic (ROC) based analysis on the MIBG features of the same clinical dataset to determine an optimal cutoff for MIBG-based diagnosis of DLB. With certain optimal cutoff value for diagnosing DLB from AD based on MIBG, an overall accuracy of 85%, a sensitivity of 71%, and specificity of 100% were obtained. In comparison, our current work (Tables II and III), using machine learning approach, had identified many possible, including alternative, assessment candidates for DLB diagnosis with accuracy ranging from 90% to 100%, sensitivity 76–100%, and specificity 88–100%. It is noteworthy that our results seemingly performed better than state-of-the-art machine learning (deep learning) approaches on neuroimaging data, which had DLB-against-AD classification accuracies in the range ∼71 – 89% [9-11].

The association rule-based classifiers derived here from the data were simple and interpretable, and hence, conducive for practical clinical use. Simple and efficient clinical tests, even tests as simple as measuring the rigidity in one limb, performed remarkably well as a diagnostic assessment feature. Specifically, the Rigid_LL feature, measuring rigidity in the left leg, which can be tested in a few seconds, was sufficient to distinguish DLB from AD in this dataset with 97% accuracy. However, it should not be assumed that such high accuracy will generalise to larger or other dataset; there is, for example, no physiological reason why the right leg rigidity should be any less predictive of DLB. Nonetheless, these results are very promising. This is particularly the case given that many different data features were selected in different rulesets, so that even if some possible candidate diagnostic tests do not generalise well to other data, it is likely that others will.

A limitation of the work is the small size of the dataset. This work uses high-dimensional data (232 features) with a small N of 33, which can create a risk of overfitting, when an algorithm optimises for patterns in the data which are not true patterns in the underlying population. This is particularly relevant when the DLB diagnosis, and thus recruitment into the original study, was based on the presence or absence of particular clinical features. Some measures used in this work to guard against overfitting included: (i) using a very simple classifier (hence with low model parameter number); (ii) using a conservative yet practical leave-one-out cross-validation technique, and testing each extracted classifier against the out-of-sample case; (iii) testing the methodology with a dataset including well-established diagnostic variables (FP-CIT) as “sanity check”; (iv) using domain knowledge to verify the extracted classifiers that seemed plausible; and (v) applying the methodology to randomly permuted data of the same dimensionality and comparing the results. Given these precautions, it is probable that many of the diagnostic rules uncovered here will generalize to other data, although the very high accuracy, sensitivity and specificity measures associated with the individual rulesets may not. The t-test performed to compare classifier error rates may also be prone to Type I error and hence, reproducibility issues.

In future work, validation and further investigation on a larger dataset is needed. A larger dataset would also allow more complex diagnostic classifiers, using more than 1-2 variables, to be extracted from the data, improving accuracy without the risk of overfitting. For example, diagnostic classifiers could be developed based on combinations of the rules extracted here. It is likely that further work on a larger dataset could develop a DLB diagnostic classifier which greatly improves on the mean out-of-sample accuracy of these classifiers (currently at 80.5%), while retaining algorithmic transparency.

Overall, while further research is needed to validate and develop the results, this work suggests there may be opportunities to refine routine clinical assessment through the introduction of quick and simple bedside tests for DLB into the dementia care pathway, thereby addressing the problem of large-scale underdiagnosis and improving outcomes and quality of life for patients with DLB.

## Data Availability

Source codes for the computations are available at https://github.com/mac-n/DLB. The dataset used during the study is available from the corresponding author (JPMK) on reasonable request.

## Appendix appendix table i

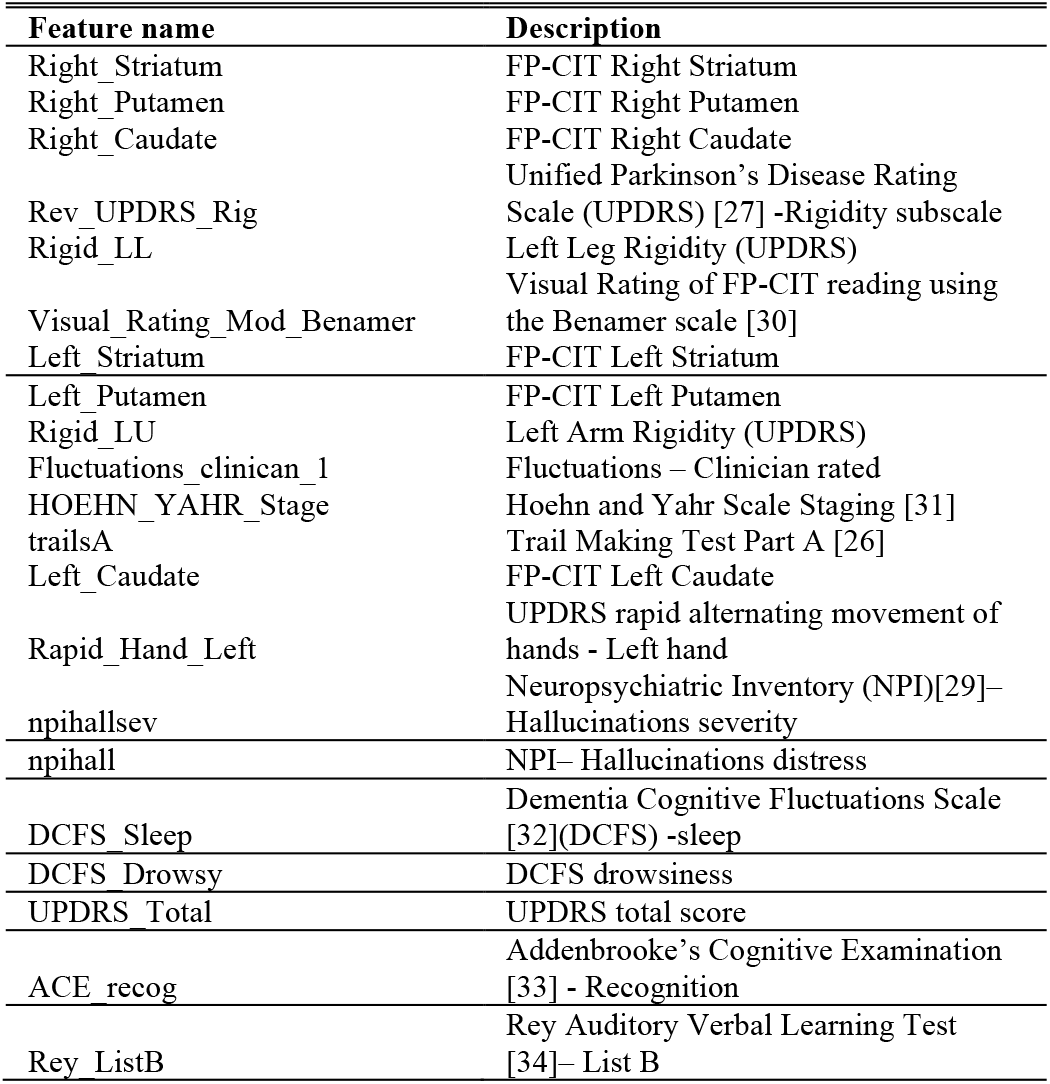
Description of Data Features Identified in this Work.

APPENDIX TABLE I DESCRIPTION OF DATA FEATURES IDENTIFIED IN THIS WORK
| Feature name | Description |
| --- | --- |
| Right_Striatum | FP-CIT Right Striatum |
| Right_Putamen | FP-CIT Right Putamen |
| Right_Caudate | FP-CIT Right Caudate |
| Rev_UPDRS_Rig | Unified Parkinson's Disease Rating Scale (UPDRS) [27] -Rigidity subscale |
| Rigid_LL | Left Leg Rigidity (UPDRS) |
| Visual_Rating_Mod_Benamer | Visual Rating of FP-CIT reading using the Benamer scale [30] |
| Left_Striatum | FP-CIT Left Striatum |
| Left_Putamen | FP-CIT Left Putamen |
| Rigid_LU | Left Arm Rigidity (UPDRS) |
| Fluctuations_clinican_1 | Fluctuations – Clinician rated |
| HOEHN_YAHR_Stage | Hoehn and Yahr Scale Staging [31] |
| trailsA | Trail Making Test Part A [26] |
| Left_Caudate | FP-CIT Left Caudate |
| Rapid_Hand_Left | UPDRS rapid alternating movement of hands - Left hand |
| npihallsev | Neuropsychiatric Inventory (NPI)[29]– Hallucinations severity |
| npihall | NPI– Hallucinations distress |
| DCFS_Sleep | Dementia Cognitive Fluctuations Scale [32](DCFS) -sleep |
| DCFS_Drowsy | DCFS drowsiness |
| UPDRS_Total | UPDRS total score |
| ACE_recog | Addenbrooke's Cognitive Examination [33] - Recognition |
| Rey_ListB | Rey Auditory Verbal Learning Test [34]– List B |

